# Using machine learning probabilities to identify effects of COVID-19

**DOI:** 10.1101/2022.07.02.22277179

**Authors:** Vijendra Ramlall, Benjamin May, Nicholas P Tatonetti

## Abstract

COVID-19, the disease caused by the SARS-CoV-2 virus, has had and continues to have extensive economic, social and public health impacts in the United States and around the world. To date, there have been more than 500 million reported cases of SARS-CoV-2 infection worldwide with more than 6 million reported deaths, more than 80 million of those cases and more than 1 million of those deaths have been reported in the United States. Retrospective analysis throughout the pandemic, which identified comorbidities, risk factors and treatments, has underpinned the response COVID-19. As the situation transitions from a pandemic to an endemic, retrospective analyses using electronic health records will be increasingly important to identify long term effects of COVID-19. However, these analyses can be complicated by the incompleteness of electronic health records, which in turns makes it difficult to differentiate visits where the patient has COVID-19. To address this, we trained a random forest classifier to assign a probability of a patient having been diagnosed with COVID-19 during each visit using demographic data, temporal data and visit-specific diagnoses (Training AUROC = 0.9867, Training OOB AUROC = 0.8957, Evaluation AUROC = 0.8958). Using these probabilities, we identified conditions associated with higher COVID-19 probabilities irrespective of clinical history and when accounting for previous diagnosis and estimated the hazards ratio for myocardial infarction (Hazards ratio = 121.736 (87.375, 169.611), *p* = 3.796E-177 and Hazards ratio = 80.262 (4.134, 4.637), *p* = 4.543E-256, respectively), urinary tract infection (Hazards ratio = 72.021 (58.116 - 89.253), *p* < 2.225E-308 and Hazards ratio = 61.380 (51.273 - 73.479), *p* < 2.225E-308, respectively), acute renal failure (Hazards ratio = 1.264E4 (9.278E4 - 1.724E4), *p* < 2.225E-308 and Hazards ratio = 6.333E3 (4.947E3 - 8.108E3), *p* < 2.225E-308, respectively) and type 2 diabetes (Hazards ratio = 345.730 (283.180 - 422.098), *p* < 2.225E-308 and Hazards ratio = 217.271 (187.898 - 251.235), *p* = 1.39E-22, respectively) when accounting for demographics and the ten most common clinical conditions.

## Introduction

The ongoing COVID-19 pandemic, caused by SARS-CoV2 infection of which there have been over 500 million cases worldwide, has resulted in more than 6.2 million deaths worldwide^1^. In the more than 30 months since the first infection is purported to have occurred^2^ and the 26 months since the start of the pandemic as declared by the World Health Organization^3^, the full impact of SARS-CoV-2 and COVID-19 remains to be seen.

Research has been paramount in responding to the COVID-19 pandemic from identifying patients susceptible to infection and at risk for severe disease^4,5,6^ to identifying beneficial treatments^7,8,9^ and developing prophylactic measures^10,11,12^. While there have been investigations into the long term effects of COVID-19^13,14,15,16,17^ continual retrospective analyses will be important to identify all the long term effects and to understand the full scope of the impact of COVID-19.

The long term effects of viral infections vary greatly. While some viruses, such as certain strains of the seasonal flu and the common cold, have no-to-little impact on the long term health of those who are infected, others can have profound long lasting effects^18,19^. Through long term analysis, it was determined that varicella zoster, the virus that causes chicken pox, also causes shingles^20^, a rash accompanied by pain, itching and tingling, in adults^21^. Retrospective analyses in patients infected with certain strains of human papilloma virus (HPV) have shown that there is an increased risk of developing anal, cervical^22,23^, penile, vaginal and vulvar cancers^24^. More recently, researchers have identified that Epstein-Barr virus, which causes mononucleosis, also triggers multiple sclerosis^25,26^, a demyelinating disease affecting the central nervous system^27^.

Much of the investigations into COVID-19, as well as varicella zoster, HPV, and Epstein-Barr virus infections, have utilized patients’ data sourced from electronic health records (EHRs). While EHRs provide a vast amount of data, such as clinical diagnoses, measurements, and procedures, they were not designed with the intention of being used for research and are incomplete. Research into COVID-19 has been further complicated by the novelty of the disease - the ICD10 code for COVID-19 (U07.1) was not effective until October 2020^28^. While the diagnosis code was indicated for COVID-19 as early as April 2020, it was not used for all COVID-19 patients nor universally adapted, which hindering differentiating COVID-19 patients from non-COVID-19 visits. To address this, we used a random forest classifier to assign a probability of a patient having had COVID-19 during each of their visits (Training Set AUROC = 0.9867, Training Set OOB AUROC = 0.8957, Evaluation Set AUROC = 0.8958).

Furthermore, we used these probabilities to identify conditions associated with a higher probability of the patient having had COVID-19 by comparing the distributions of COVID-19 probability of visits that were followed with the diagnosis of a conditions at 1 week, 2 weeks, 3 weeks, 4 weeks, 3 months, 6 months, 9 months and 1year using a Mann-Whitney U test. In applying a Cox proportional hazards model, we identified myocardial infarction ((Hazards ratio = 121.736 (87.375, 169.611), *p* = 3.796E-177 and Hazards ratio = 80.262 (4.134, 4.637), *p =* 4.543E-256, respectively), urinary tract infection (Hazards ratio = 72.021 (58.116 - 89.253), *p* < 2.225E-308 and Hazards ratio = 61.380 (51.273 - 73.479), *p* < 2.225E-308, respectively), acute renal failure (Hazards ratio = 1.264E4 (9.278E4 - 1.724E4), *p* < 2.225E-308 and Hazards ratio = 6.333E3 (4.947E3 - 8.108E3), *p* < 2.225E-308, respectively) and type 2 diabetes (Hazards ratio = 345.730 (283.180 - 422.098), *p* < 2.225E-308 and Hazards ratio = 217.271 (187.898 - 251.235), *p* = 1.39E-22, respectively) when accounting for demographics and the ten most common clinical conditions.

## Results

From the clinical data at New York-Presbyterian, we identified 1,844,018 visits for 636,063 patients who sought treatment at least once between February 1st, 2020 and March 31st, 2022 at /Columbia University Irving Medical Center (NYP/CUIMC). We omitted 270,905 visits for 201,911 patients who did not not have any demographic data available in our clinical data set (Figure 1). From these visits, we identified 9,340 visits (COVID-19 visits) where the patient was diagnosed with COVID-19 evidenced by the presence of the COVID-19 ICD-10 diagnosis code (U07.1) (Figure 1). Additionally, we identified 1,483,397 visits (non-COVID-19 visits) where the patient did not test positive for SARS-CoV-2 during that visit nor had a history of COVID-19 nor previously tested positive for SARS-CoV-2 infection (Figure 1). The set of COVID-19 visit was randomly split into distinct testing and evaluation sets, each with 4,670 visits and from the set of non-COVID-19 visits, we randomly identified distinct testing and evaluation non-COVID-19 sets, each with 4,670 unique visits.

**Figure 1.**
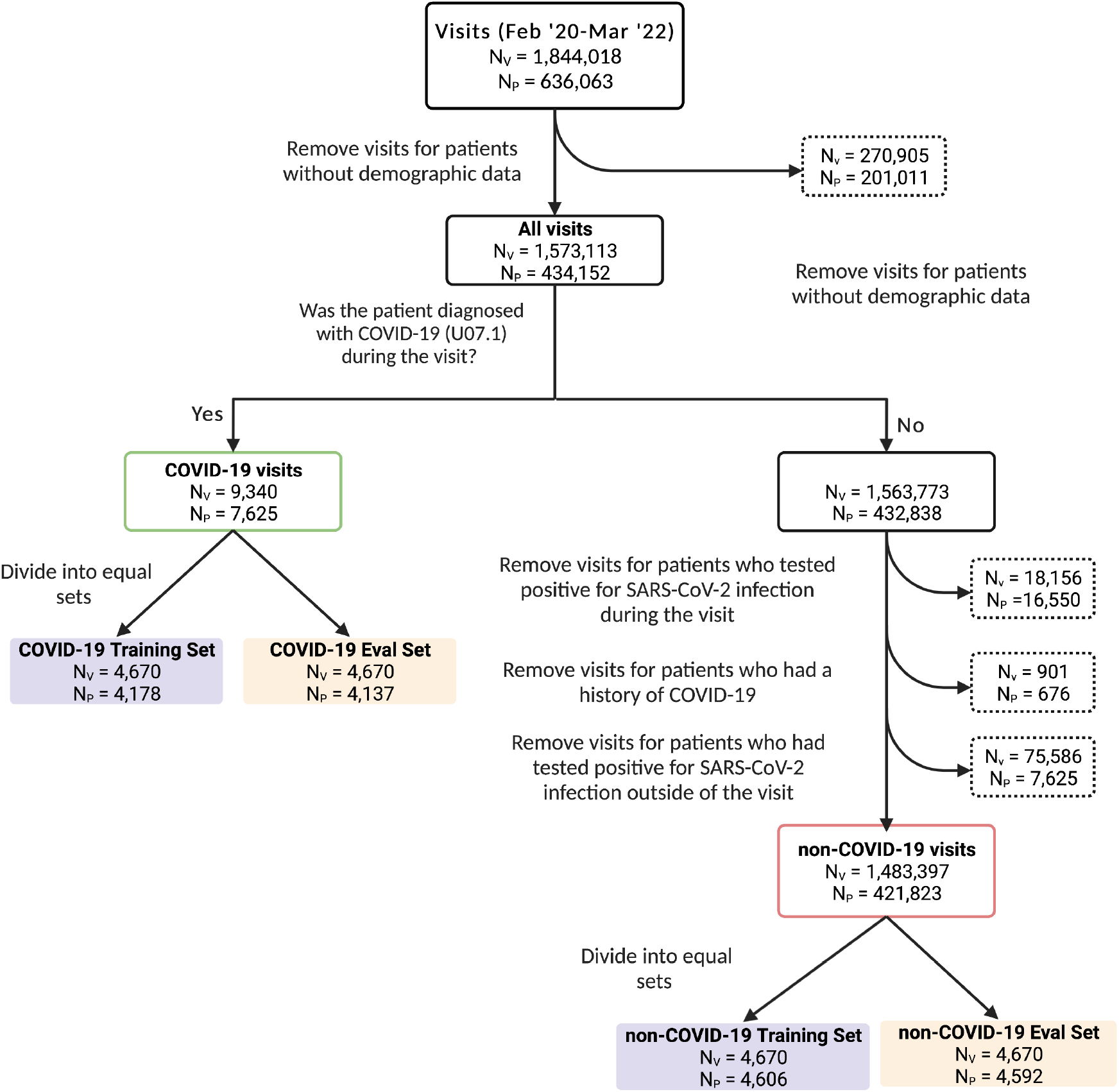
Data processing flowchart. Identification of COVID-19 and non-COVID-19 training sets (purple) and evaluation sets (orange). N_V_ indicates the number of visits and N_P_ indicates the number of patients in each group. Note: the exclusion criteria used to identify non-COVID-19 visits are not mutually exclusive.

Among all visits between February 2020 and March 2022, as well as the COVID-19 and non-COVID-19 training and evaluations sets, more than 50% of the visits were for patients who self identified as female and more than 85% of the visits were for patents who were at least 19 years old (adults and senior age groups) (Table 1). Across all of the groups, more than 35% of the visits were for patients who self identified as White, more than 15% were for patients who self identified as Black or African American and more than 29% were visits for patients who self identified as Hispanic or of Latino or Spanish origin (Table 1). In all groups, less than 5% of visits were for patients who self identified as American Indian or Alaskan Native, Asian or Native Hawaiian or Other Pacific Islander (Table 1).

**Table 1.** Demographics of patients of visits used for model training, model evaluation and all visits between February 2020 and March 2022.

|  | Model Training Set |  | Model Evaluation Set |  | All Visits |
| --- | --- | --- | --- | --- | --- |
|  | non-COVID-19 | COVID-19 | non-COVID-19 | COVID-19 | Feb 2020 - Mar 2022 |
| **N(visits)** | 4,670 | 4,670 | 4,670 | 4,670 | 1,573,113 |
| **N(patients)** | 4,606 | 4,178 | 4,592 | 4,137 | 434,152 |
| **Age Child (< 13)** | 365 | 235 | 326 | 242 | 122,140 |
| **(% of visits)** | 7.82% | 5.03% | 6.98% | 5.18% | 7.76% |
| **Age Adolescent (≥ 13 and < 19)** | 146 | 115 | 156 | 115 | 49,232 |
| **(% of visits)** | 3.13% | 2.46% | 3.34% | 2.46% | 3.13% |
| **Age Adult (≥ 19 and < 60)** | 2,375 | 2,159 | 2,324 | 2,099 | 772,606 |
| **(% of visits)** | 50.86% | 46.23% | 49.76% | 44.95% | 49.11% |
| **Age Senior (≥ 60)** | 1,784 | 2,161 | 1,864 | 2,214 | 629,135 |
| **(% of visits)** | 38.20% | 46.27% | 39.91% | 47.41% | 39.99% |
| **Self Identified Sex as Female** | 2,756 | 2,412 | 2,885 | 2,403 | 941,558 |
| **(% of visits)** | 59.01% | 51.65% | 61.78% | 51.46% | 59.85% |
| **Self Identified as American Indian or Alaskan Native** | 14 | 18 | 11 | < 10 | 3,994 |
| **(% of visits)** | 0.30% | 0.39% | 0.24% | < 0.41% | 0.25% |
| **Self Identified as Asian** | 108 | 124 | 120 | 116 | 39,091 |
| **(% of visits)** | 2.31% | 2.66% | 2.57% | 2.48% | 2.48% |
| **Self Identified as Black or African American** | 728 | 803 | 702 | 811 | 245,104 |
| **(% of visits)** | 15.59% | 17.19% | 15.03% | 17.37% | 15.58% |
| **Self Identified as Native Hawaiian or Other Pacific Islander** | < 10 | 10 | < 10 | < 10 | 1,520 |
| **(% of visits)** | < 0.21% | 0.21% | < 0.21% | < 0.21% | 0.10% |
| **Self Identified as White** | 1,900 | 1,645 | 1,974 | 1,669 | 643,848 |
| **(% of visits)** | 40.69% | 35.22% | 42.27% | 35.74% | 40.93% |
| **Self identified as Hispanic or of Latino or Spanish Origin** | 1,433 | 1,873 | 1,367 | 1,782 | 473,501 |
| **(% of visits)** | 30.69% | 40.11% | 29.27% | 38.16% | 30.10% |

Among all visits between February 2020 and March 2022, the largest fraction of visits (5.17%) began in March 2021 (Table S1). The largest fraction of visits in the COVID-19 training and evaluation sets began in April 2020 (18.29% and 17.99%, respectively), while the smallest fraction of all visits began in April 2020 until March 2022 (1.64%) (Table S1). The fraction of visits in the non-COVID-19 training and evaluation sets that began in each month were similar to the faction of all visits that began in each month (Table S1). Among all visits between February 2020 and March 2022, the four diagnosis listed in the most visits were encounter for supervision of normal pregnancy (2.38%), transplanted organ and tissue status (2.26%), other symptoms and signs involving the circulatory and respiratory system (2.18%) and essential (primary) hypertension (2.06%) (Table S2). Among the COVID-19 visits in the training and evaluation sets, diagnosis of other symptoms and signs involving the circulatory and respiratory system (20.75% and 19.21%, respectively), encounter for screening for malignant neoplasms (19.46% and 19.08%, respectively), essential (primary) hypertension (8.84% and 9.27%, respectively) and transplanted organ and tissue status (8.22% and 8.78%, respectively) were frequently diagnosed (Table S2). The fraction of non-COVID-19 visits in the training and evaluation sets with the diagnoses listed was similar to the fraction of all visits with the diagnosis listed (Table S2).

We collected demographic data for the patient in each visit (date of birth, self-identified sex, self-identified race(s) and self-identified ethnicity), temporal data (during what month the visit started) and visit specific diagnosis data. In our dataset, there were 16,220 distinct ICD10 codes used to records diagnoses which we generalized to 1,600 category level ICD10 codes. We decided to use a random forest classifier to predict whether or not a patient was diagnosed with COVID-19 during their visit using demographic, temporal, and visit-specific clinical diagnoses. The diagnosis code for COVID-19 (U07.1) was removed from the data to be used in the training the model prior to generalization. Instead of binary outcome (patient having been diagnosed with COVID-19 during their visit or not), we used the fraction of estimators identifying the visit as one where the patient was diagnosed with COVID-19 as the probability of the patient having COVID-19 during the visit. An initial random forest classifier of 200 estimators was fit using the COVID-19 and non-COVID-19 training sets with bootstrapped sampling and using out-of-bag sampling (Training AUROC = 0.9923, Training OOB AUROC = 0.8838, Evaluation AUROC = 0.8838) (Figure S1A). In order to optimize the performance of the model, we monitored the AUROC of the training set, the training set using out-of-bag estimates and the evaluation set while increasing the number of estimators from 20 to 200 and achieved a maximum AUROC in the evaluation set with 190 estimators (Training Set AUROC = 0.9924, Training Set OOB AUROC = 0.8836, Evaluation Set AUROC = 0.8839) (Figure 2A). We further optimized the performance of the model by monitoring the AUROC while increasing the maximum depth of the model from 1 to 100 with 190 estimators and achieved a maximum AUROC in the evaluation set with a depth of 69 (Training Set AUROC = 0.9867, Training Set OOB AUROC = 0.8957, Evaluation Set AUROC = 0.8958) (Figure 2B). The optimized model trained with 190 estimators with a maximum depth of 69 was fit to the data representing all 1,573,113 visits (Figure 2C).

**Figure 2.**
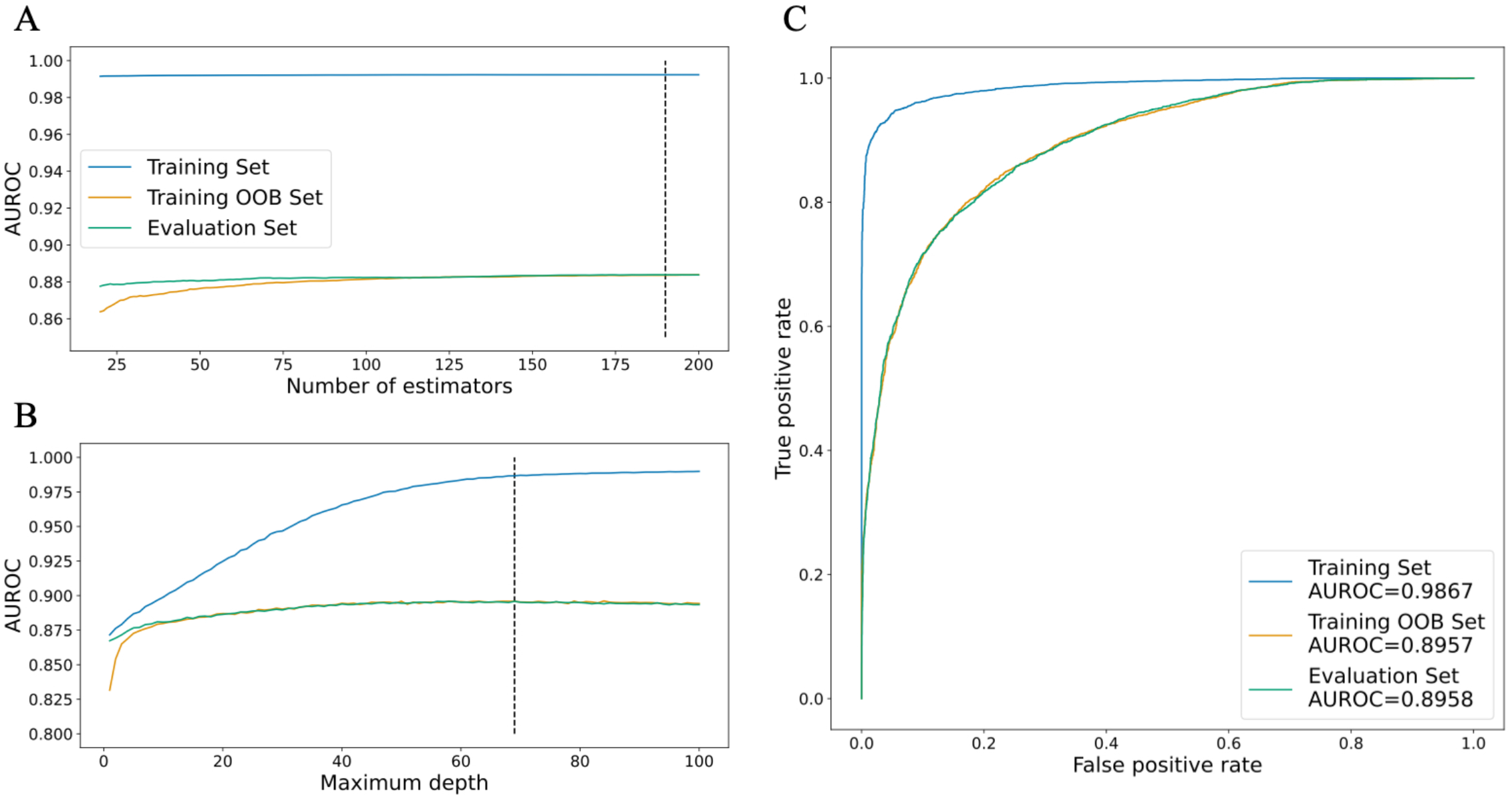
Model performance optimization. (A) AUROC in training set, training set using out-of-bag estimates, and evaluation set plotted against number of estimators (dashed line indicates maximum AUROC in evaluation set, n_estimators = 190) (B) AUROC in training set, training set using out-of-bag estimates, and evaluation set plotted against maximum depth (dashed line indicates maximum AUROC in evaluation set, max_depth = 69) (C) ROC curves of training set, training set using out-of-bag estimates, and evaluation set.

We evaluated the features utilized in the final model using the Gini importance (Table 2, Table S3). Diagnosis of abnormalities of breathing (R06), other symptoms and signs involving the circulatory and respiratory system (R09) and cough (R05) during the visit had the highest importance in the final model (Table 2). The distribution of the COVID-19 probabilities of the visits where the diagnoses were noted were skewed to higher COVID-19 probability than those where the diagnosis were not noted in both the training and evaluation sets (Wasserstein distance = 0.4602, 4510, 0.4458, respectively in the training set) (Figure 3 B-D, Table 2). Visits starting in April 2020, June 2021 and July 2021 were the temporal features with the highest importance in the final model (Table 2). The distribution of the COVID-19 probabilities of visits that started in April 2020 were skewed to higher COVID-19 probabilities than those that did not start in April 2020 (Wasserstein distance = 0.4353 in the training set) (Figure 3E, Table 2). Conversely, the distributions of the COVID-19 probabilities of visits that started in June 2021 and July 2021 were skewed to lower COVID-19 probabilities than those started at other times (Wasserstein distance = 0.3871, 0.3780, respectively in the training set) (Figure 3F-G, Table 2). Patients self-identifying as White, of Hispanic or Latino or Spanish origin, and female were the demographic features with the highest importance in the final model (Table 2). The distributions of COVID-19 probabilities of visits where the patients self identified as White or female were skewed to lower COVID-19 probabilities that those where the patient did not (Wasserstein distance = 0.0573, 0.0711, respectively in the training set) (Figure 3H, 3J, Table 2). The distribution of COVID-19 probabilities of visits where the patients self identified as of Hispanic or Latino or Spanish origin were skewed to higher COVID-19 probabilities that those where the patient did not (Wasserstein distance = 0.0920) (Figure 3I, Table 2).

**Table 2.** Importance for the top 20 important features and Wasserstein distance between distribution where the feature is observed and the feature is not observed. Negative Wasserstein distance indicates that the average COVID-19 probability in the set of visits where the feature was observed is less than the average of the set where the feature was not observed.

| Feature | Importance | Wasserstein Distance |  |  |
| --- | --- | --- | --- | --- |
|  |  | Training Set | Evaluation Set | All Visits |
| Abnormalities of breathing diagnosis noted during visit (R06) | 0.0650 | 0.4602 | 0.4640 | 0.5074 |
| Other symptoms and signs involving the circulatory and respiratory system diagnosis noted during visit (R09) | 0.0628 | 0.4510 | 0.4355 | 0.4718 |
| Visit started in April 2020 | 0.0543 | 0.4353 | 0.4371 | 0.4209 |
| Cough diagnosis noted during visit (R05) | 0.0259 | 0.4458 | 0.4293 | 0.5160 |
| Viral pneumonia, not elsewhere classified diagnosis noted during visit (J12) | 0.0236 | 0.4979 | 0.4777 | 0.6738 |
| Encounter for other special examination without complaint, suspected or reported diagnosis diagnosis noted during visit (Z01) | 0.0234 | 0.2969 | 0.3009 | 0.4270 |
| Transplanted organ and tissue status diagnosis noted during visit (Z94) | 0.0229 | 0.3066 | 0.3145 | 0.4175 |
| Fever of other and unknown origin diagnosis noted during visit (R50) | 0.0195 | 0.4400 | 0.4169 | 0.5089 |
| Respiratory failure, not elsewhere classified diagnosis noted during visit (J96) | 0.0176 | 0.5022 | 0.4920 | 0.6076 |
| Self Identified as White | 0.0148 | -0.0573 | -0.0615 | -0.0079 |
| Visit started in June 2021 | 0.0148 | -0.3871 | -0.3630 | -0.2007 |
| Self identified as of Hispanic or Latino or Spanish Origin | 0.0141 | 0.0920 | 0.0899 | 0.0287 |
| Self Identified Sex as Female | 0.0141 | -0.0711 | -0.0759 | -0.0187 |
| Visit started in July 2021 | 0.0130 | -0.3780 | -0.3570 | -0.1868 |
| Visit started in August 2021 | 0.0126 | -0.3432 | -0.3643 | -0.1722 |
| Visit started in September 2021 | 0.0117 | -0.3439 | -0.3266 | -0.1826 |
| Visit started in February 2020 | 0.0115 | -0.3903 | -0.3221 | -0.1221 |
| Visit started in October 2021 | 0.0112 | -0.3539 | -0.3586 | -0.1855 |
| Type 2 diabetes mellitus diagnosis noted during visit (E11) | 0.0106 | 0.4029 | 0.3949 | 0.4329 |
| Acute kidney failure diagnosis noted during visit (N17) | 0.0104 | 0.4751 | 0.4700 | 0.5350 |

**Figure 3.**
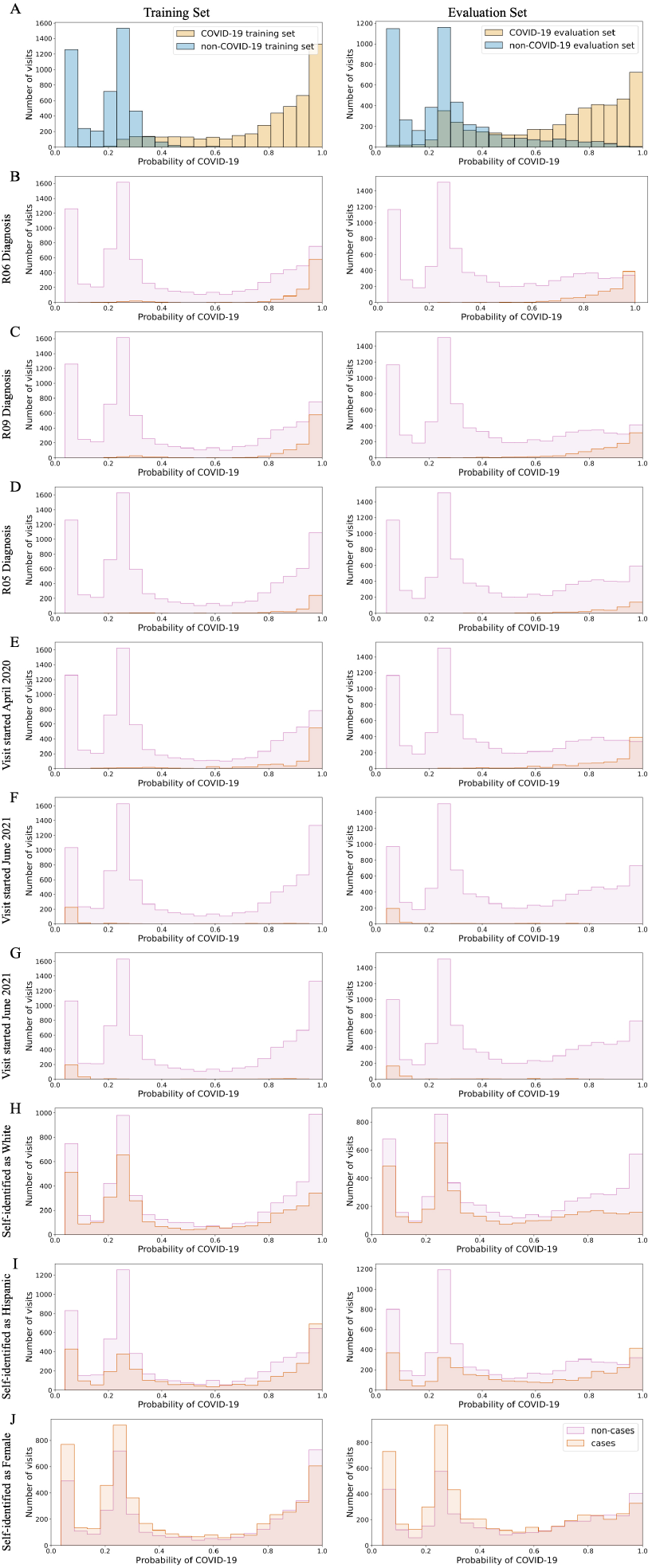
Distribution important features in random forest classifier in training and evaluation sets. (A) Distribution of COVID-19 probability in COVID-19 (yellow) and non-COVID-19 (blue) training (left) and evaluations (rights) sets (top). Distribution of cases (red) and non-cases (purple) for important diagnoses (B-D), temporal (E-G) and demographic (H-J) features for training and evaluation sets. Note: R06 - abnormalities of breathing, R09 - other symptoms and signs involving the circulatory and respiratory system diagnosis noted during visit, R05 - cough.

We further evaluated the model by evaluating the distributions of COVID-19 probabilities for visits within inclusion and exclusion criteria for the training and evaluation sets (Figure 1). Compared to the distribution of COVID-19 probabilities for all of the visits between February 2020 and March 2022 (Figure 4A), visits where the patient was diagnosed with COVID-19 based on the presence of the U07.1 ICD-10 code (N=9,340) during the visits were skewed to higher COVID-19 probabilities (Wasserstein distance = 0.4695) (Figure 4B). The distribution of COVID-19 probabilities of visits where the patient tested positive for SARS-CoV-2 infection (N=18,156) was bimodal with a skewed to higher COVID-19 probabilities (Wasserstein distance = 0.2319) (Figure 4C). The distribution of COVID-19 probabilities of visits where the patient tested negative for SARS-CoV-2 infection (N=238,438) was marginally skewed to to higher COVID-19 probabilities (Wasserstein distance = 0.0550) (Figure 4D). The distribution of COVID-19 probabilities of visits where clinical diagnosis notes indicated that the patient did not have COVID-19 (N=168) was skewed to higher COVID-19 probabilities (Wasserstein distance = 0.4158) (Figure 4E).The distribution of COVID-19 probabilities of visits where the patient had a noted history of COVID-19 (N=899) was skewed to higher COVID-19 probabilities (Wasserstein distance = 0.3547) (Figure 4F).

**Figure 4.**
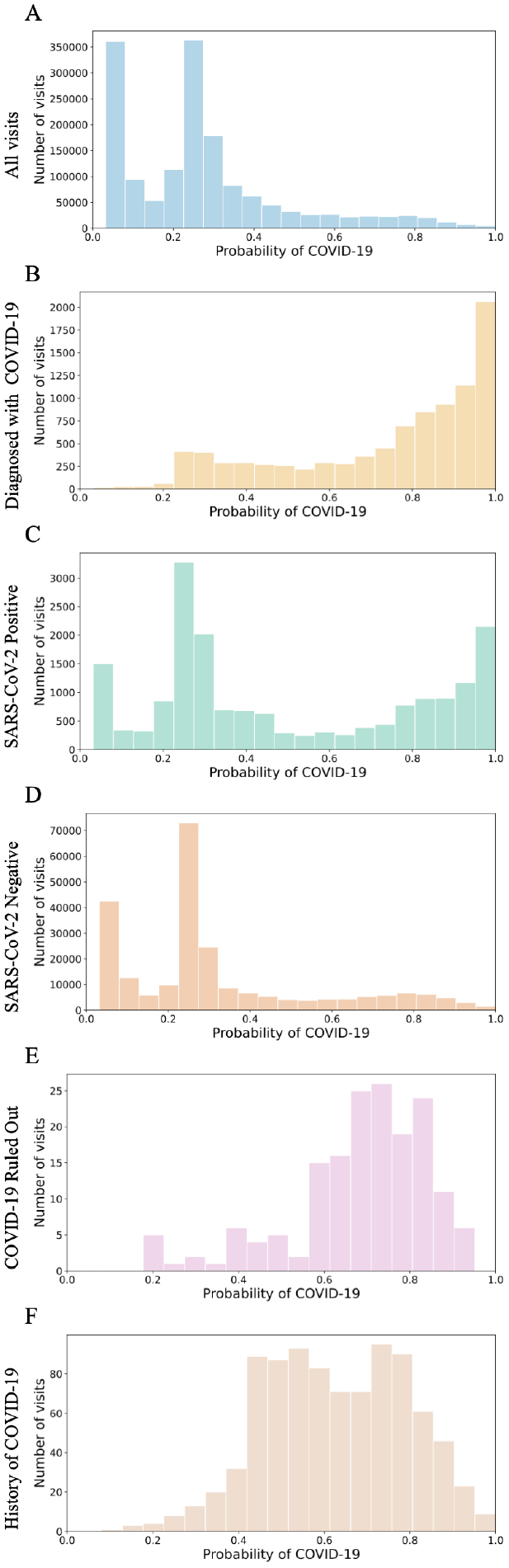
Distribution of COVID-19 probability for visits different patient groups. (A) Distribution of COVID-19 probability for all visits. Distribution of visits where patients were diagnosed with COVID-19 (B), tested positive for SARS-CoV-2 infection (C), tested negative for SARS-CoV-2 infection (D), where clinical diagnosis note indicated the “COVID-19 was ruled out” (E) and visits where the patient had a history of COVID-19 (F).

In order to identify what, if any, conditions are associated with a history COVID-19, we identified visits where the patient returned to the hospital within 7 days, 14 days, 21 days, 28 days, 3 months, 6 months, 9 months and 12 months by comparing the distributions of COVID-19 probabilities of visits where the patient returned within each time period and then segregated the visits into those where a particular condition was observed in the followup and those where the condition was not (Figure 1). We used a Mann-Whitney U test to compare between the two distributions for each conditions irrespective of whether or not the patient was previously diagnosed with the condition (Figures 5 left, Table S4) and only if the patient was not diagnosed with the condition prior to the visit (Figure 5 right, Table S5). We identified, among other conditions, the distribution of COVID-19 probability preceding myocardial infarction was significantly different from the distribution of COVID-19 probability not preceding myocardial infarction both with and without accounting for previous clinical history in all time periods (Mann-Whitney U test statistic = 1.206E8, FDR correct *p* < 2.225E-308, Mann-Whitney U test statistic = 1.339E8, FDR correct *p* < 2.225E-308, respectively within one year) (Figure 5). We observed a similar difference with and without accounting for previous clinical history for urinary tract infection (Mann-Whitney U test statistic = 1.968E8, FDR correct *p* < 2.225E-308, Mann-Whitney U test statistic = 2.562E8, FDR correct *p* < 2.225E-308 within one year), acute renal failure (Mann-Whitney U test statistic = 8.969E7, FDR correct *p* < 2.225E-308, Mann-Whitney U test statistic = 1.234E8, FDR correct *p* < 2.225E-308 within one year), and type 2 diabetes (Mann-Whitney U test statistic = 2.317E8, FDR correct *p* < 2.225E-308, Mann-Whitney U test statistic = 3.273E8, FDR correct *p* < 2.225E-308 within one year) (Figure 5).

**Figure 5.**
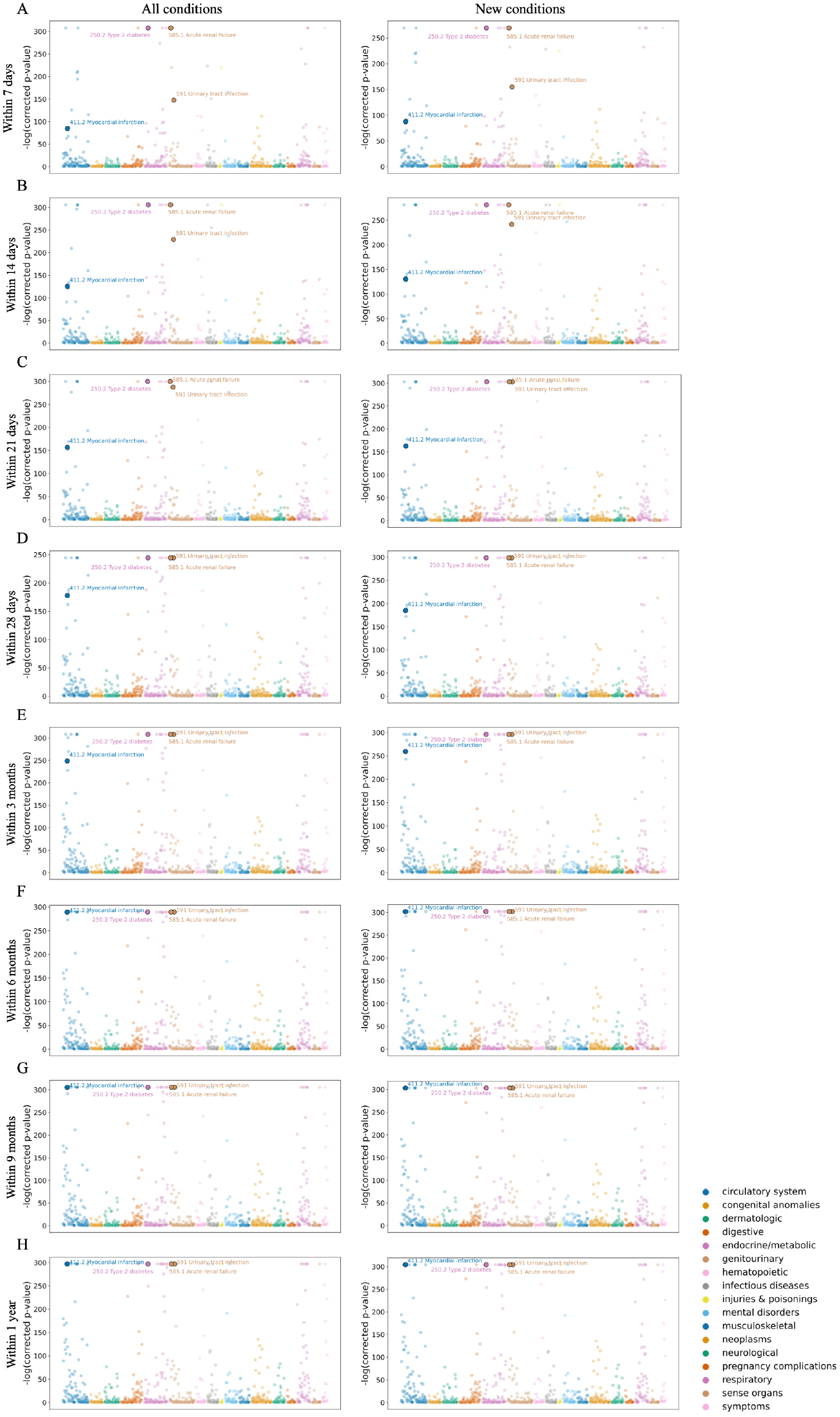
Statistical testing of conditions associated with COVID-19. -log_10_(corrected *p*-value) for each phenotype (colored by family) from Mann-Whitney U test between distributions of COVID-19 probabilities of cases and non-cases for each phenotype within (A) 7 days, (B) 14 days, (C) 21 days, (D) 28 days, (E) 3 months, (F) 6 months, (G) 9 months and (H) 1 year irrespective of previous clinical list (left) and when accounting for clinical history (right).

To further investigate the association between COVID-19 probability and the onset of conditions, we calculated the hazard ratio using a Cox proportional hazards model for COVID-19 probability irrespective of previous clinical history (Figure 6 A left) and respective of previous clinical history (Figure 6 A right). Increasing COVID-19 probability in the preceding visit was associated with increase risk of myocardial infarction within one year with and without accounting for previous clinical history (Hazards ratio = 93.713 (73.906-118.829), *p* = 2.199E-307 and Hazards ratio = 82.557 (65.102-104.693), *p* = 2.414E-290, respectively) (Table 3). A similar association was observed within one year with and without accounting for previous clinical history for urinary tract infection (Hazards ratio = 75.241 (63.192 - 89.587), *p* < 2.225E-308 and Hazards ratio = 62.038 (52.176 -73.765), *p* < 2.225E-308, respectively), acute renal failure (Hazards ratio = 7762.722 (6156.997 - 9787.216), *p* < 2.225E-308 and Hazards ratio = 5488.974 (4345.262 - 6933.722), *p* < 2.225E-308, respectively) and type 2 diabetes (Hazards ratio = 403.553 (350.901 - 464.106), *p* < 2.225E-308 and Hazards ratio = 270.035 (235.213 - 310.013), *p* < 2.225E-308, respectively) (Table 3).

**Figure 6.**
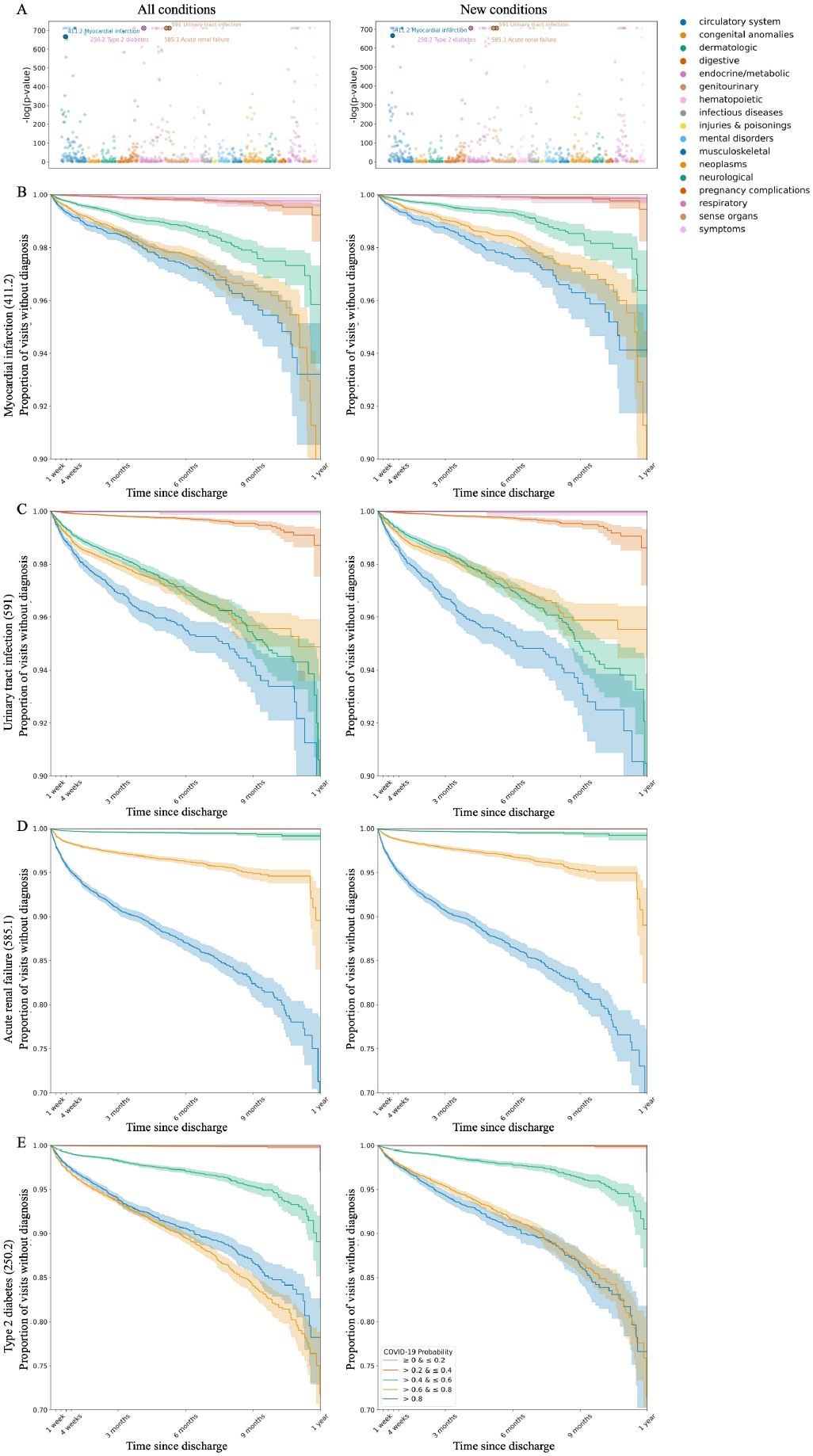
Statistical testing of conditions associated with COVID-19. (A)-log_10_(*p*-value) for each phenotype (colored by family) from Cox Proportional Hazards test for COVID-19 probability of the previous visit for conditions developed within 1 year irrespective of clinical history (left) and when accounting for clinical history (right). Kaplan-Meier curves for (B) myocardial infarction, (C) urinary tract infection, (D) acute renal failure, (E) type 2 diabetes stratified by COVID-19 probability quintile within 1 year irrespective of clinical history (left) and when accounting for clinical history (right).

**Table 3.** Results of Mann-Whitney U test, univariate Cox proportional hazards ratio and multivariate Cox proportional hazards ratio for COVID-19 probability within 1 year.

|  | Myocardial infarction (411.2) |  | Urinary tract infection (591) |  | Acute renal failure (585.1) |  | Type 2 diabetes (250.2) |  |
| --- | --- | --- | --- | --- | --- | --- | --- | --- |
|  | All conditions | New Conditions | All conditions | New Conditions | All conditions | New Conditions | All conditions | New Conditions |
| **Mann-Whitney U Test** | 1.339E+08 | 1.206E+08 | 2.562E+08 | 1.968E+08 | 1.234E+08 | 8.969E+07 | 3.273E+08 | 2.317E+08 |
| **(Test statistic, *p* value, FDR** | 2.944E-299 | 1.793E-306 | 2.944E-299 | 1.793E-306 | 2.944E-299 | 1.793E-306 | 2.944E-299 | 1.793E-306 |
| **corrected *p* value** | < 2.225E-308 | < 2.225E-308 | < 2.225E-308 | < 2.225E-308 | < 2.225E-308 | < 2.225E-308 | < 2.225E-308 | < 2.225E-308 |
| **Cox Proportional Hazards** | 82.557 | 93.713 | 62.038 | 75.241 | 5488.974 | 7762.723 | 270.035 | 403.553 |
| **Univariate Fit** | (65.102, 104.693) | (73.906, 118.829) | (52.176, 73.765) | (63.192, 89.587) | (4.345E3, 6.934E3) | (6.157E3, 9.787E3) | (235.213, 310.013) | (350.901, 464.106) |
| **(Hazards ratio, 95% CI, *p* value)** | 2.414E-290 | 2.199E-307 | < 2.225E-308 | < 2.225E-308 | < 2.225E-308 | < 2.225E-308 | < 2.225E-308 | < 2.225E-308 |
| **Cox Proportional Hazards** | 80.262 | 121.736 | 61.380 | 72.021 | 6333.163 | 12647.836 | 217.271 | 345.730 |
| **Multivariate Fit** | (62.417, 103.208) | (87.375, 169.611) | (51.273, 73.479) | (58.116, 89.253) | (4.947E3, 8.108E3) | (9.278E3, 1.724E4) | (187.898, 251.235) | (283.180, 422.098) |
| **(Hazards ratio, 95% CI, *p* value)** | 4.543E-256 | 3.796E-177 | < 2.225E-308 | < 2.225E-308 | < 2.225E-308 | < 2.225E-308 | < 2.225E-308 | < 2.225E-308 |

Among the visits with a followup within one year, the ten most frequently observed phenotypes were essential hypertension (401.1), shortness of breath (512.7), hyperlipidemia (272.1), other complications of pregnancy NEC (646), cough (512.8), back pain (760), injury, NOS (1009), gastroesophageal reflux disease (530.11), other headache syndromes (339), and pulmonary collapse; interstitial and compensatory emphysema (508), respectively. When accounting for demographics and the ten most frequently observed phenotypes in a multivariate Cox proportional hazards model, increasing COVID-19 probability in the preceding visit was associated with increase risk of myocardial infarction within one year with and without accounting for previous clinical history (Hazards ratio = 121.736 (87.375, 169.611), *p* = 3.796E-177 and Hazards ratio = 80.262 (4.134, 4.637), *p* = 4.543E-256, respectively) (Table 3). A similar association was observed within one year with and without accounting for previous clinical history for urinary tract infection (Hazards ratio = 72.021 (58.116 - 89.253), *p* < 2.225E-308 and Hazards ratio = 61.380 (51.273 - 73.479), *p* < 2.225E-308, respectively), acute renal failure (Hazards ratio = 1.264E4 (9.278E4 - 1.724E4), *p* < 2.225E-308 and Hazards ratio = 6.333E3 (4.947E3 - 8.108E3), *p* < 2.225E-308, respectively) and type 2 diabetes (Hazards ratio = 345.730 (283.180 - 422.098), *p* < 2.225E-308 and Hazards ratio = 217.271 (187.898 - 251.235), *p* = 1.39E-22, respectively) (Table 3, S6).

We further stratified the COVID-19 probabilities into quintiles and generated Kaplan-Meier curves for the data within one year (Figure 6B-E). The Kaplan-Meier curves stratified by COVID-19 probability for myocardial infarction showed three distinct sets, (i) COVID-19 probability greater than 0.6, (ii) COVID-19 probability greater than 0.4 and less than or equal to 0.6 and (iii) COVID-19 probability less than or equal to 0.4, with higher incidence observed in the sets of higher COVID-19 probability (Figure 6B). The Kaplan-Meier curves for urinary tract infection showed three sets, (i) COVID-19 probability greater than 0.8, (ii) COVID-19 probability greater than 0.4 and less than or equal to 0.8 and (iii) COVID-19 probability less than or equal to 0.4, up to 8 months with the higher incidence observed in the sets of higher COVID-19 probability (Figure 6C). The Kaplan-Meier curves for acute renal failure showed four distinct sets, (i) COVID-19 probability greater than 0.8, (ii) COVID-19 probability greater than 0.6 and less than or equal to 0.8, (iii) COVID-19 probability greater than 0.4 and less than or equal to 0.6 and (iii) COVID-19 probability less than or equal to 0.4, with the higher incidence observed in the sets of higher COVID-19 probability (Figure 6D). The Kaplan-Meier curves for the onset of type 2 diabetes showed three distinct sets (i) COVID-19 probability greater than 0.6, (ii) COVID-19 probability greater than 0.4 and less than or equal to 0.6 and (iii) COVID-19 probability less than or equal to 0.4, with higher incidence observed in the sets of higher COVID-19 probability (Figure 6E).

## Discussion

In this study, we collected demographic, temporal and clinical data from 434,152 patients who sought treatment at New York-Presbyterian over 1,573,113 visits between February 2020 and March 2022, who had at least one interaction with Columbia University Irving Medical Center, to develop an algorithm to identify conditions that are associated with COVID-19. The 26 month period from which our data is sourced encompasses the height of the first wave of the COVID-19 pandemic (Spring 2020) when New York City was an epicenter in the United States as well as the subsequent Delta and Omicron waves^29^. Additionally, our data encompasses periods, such as summer 2020 when case counts were at some of their lowest levels throughout the pandemic, as well as the period following development of treatments for COVID-19 and prophylactics for SARS-CoV-2 infection.

Using data for patients who had COVID-19 diagnosed (as determined by the presence of the U07.1 ICD-10 diagnosis code) and non-COVID-19 patients, we trained an optimized random forest classifier with high performance as evaluated in an independent data set, and applied it to the full set of 1,573,113 visits. Instead of the binary classification that would result from the random forest classifier, we instead treated the fraction of estimators that identified the visit as a COVID-19 visit as a probability of a patient having been diagnosed with COVID-19 during that visit. While the random forest classifier is overfitting based on the high AUROC observed in the training set, we were comfortable using it because it performed similarly in the training set using out-of-bag estimates and the evaluation set. Based on the presence of U07.1 ICD-10 diagnosis code, there were only 9,340 where the patient was diagnosed with COVID-19, however our model identified 198,562 visits where the patients had a probability of having been diagnosed with COVID-19 greater than 0.5.

When evaluating our model, the most important features represented previously identified differences between demographic groups, such as those who identify as Hispanic or Latino or of Spanish origin or Black or African American^30,31^ (Table 2, Figure 3). Important temporal features represented periods of extreme case counts in New York City, such as spring 2020 and summer 2021^27^ (Table 2, Figure 3). Important clinical diagnoses were reflective of known symptoms of COVID-19^32^, such as abnormalities of breathing (R06), other symptoms and signs involving the circulatory and respiratory system (R09) and cough (R05) (Table 2, Figure 3).

Using these visit specific probabilities, we identified conditions that developed within different time periods after the visit (up to 7 days, 14 days, 21 days, 28 days, 3 moths, 6 months, 9 months, and 12 months) and used a Mann-Whitney U test to identify conditions that were associated with increased COVID-19 probability. Among others, our analysis identified myocardial infarction, urinary tract infection, acute renal failure and type 2 diabetes as being associated with. COVID-19 (Figure 5). In further analysis of the results of our results, we estimated the hazards ratio of COVID-19 probability for each of these conditions (Table 3). Cox proportional hazards model indicated that higher COVID-19 probability in the preceding visit was associated with an increased risk of myocardial infarction, urinary tract infection, acute renal failure and type 2 diabetes within one year. Our result for myocardial infarction is consistent with those of researchers who identified a higher risk of heart attack and ischemic stroke in COVID-19 patients using self-controlled case series^14^. Results from a retrospective observational study of patients in early 2020 observed that severe COVID-19 disease is associate with acute kidney injury^16^. Other researchers have identified an increased risk of type 2 diabetes in patients who had been infected with SARS-CoV-2 compared to patient who had not and compared to a historical control^17^.

While this study shows that demographic, temporal and clinical data can be utilized to predict the probability of a patient having COVID-19 during their visit, the model and the important features are specific to NYP/CUIMC. An implementation this model elsewhere is expected to identify important temporal features specific to the site (e.g. periods of extreme case counts varied between New York City and London) and demographic variables depending on the patients seeking treatment at those sites. However, it would be expected to identify similar clinical variables that are representative of known symptoms or comorbidities associated with COVID-19. While the results concur with other studies, they are not without their biases as this study relied on patients who sought treatment at New York-Presbyterian on multiple occasions and was unable to incorporate data from patients who may have also sought outside treatment due to the nature of primary care in the United States. Finally, in identifying effects of COVID-19, we are limited by the novelty of the disease itself since other effects may take years or decades to develop.

## Conclusion

Our study demonstrated a new method to conduct retrospective analyses for identifying the effects of COVID-19. By implementing a model trained on clinical data at the visit level and using the output from a random forest classifier as a probability instead of a binary outcome, we mitigated the need to definitively distinguish cases. Additionally, the results from our study can be used to direct further investigations into the effects of COVID-19. As the COVID-19 pandemic transitions to an endemic situation, our method can be utilized to understand potential pathophysiological difference in symptoms associated with COVID-19 spikes. Moreover, as this method was designed using concurrent clinical data, it can be adapted to other novel or emerging diseases.

## Methods

### Ethics statement

The study is approved by the Columbia University Irving Medical Center Institutional Review Board (IRB) no. AAAL0601 and the requirement for informed consent was waived. A data request associated with this protocol was submitted to the Tri-Institutional Request Assessment Committee of New York-Presbyterian/Columbia and Cornell and approved.

### Preparation for data modeling and statistical modeling

We used MySQL 5.7.35 and Python 3.9.10 with numpy 1.19.5, pymysql 1.0.2, and pandas 1.2.3 libraries to extract and prepare data for modeling. For each visit, we identified the age of the patient at the start of the visit as (i) birth to 13 years old, (ii) 13 to 19 years old, (iii) 19 to 60 years old and (iv) over 60 years old and if the patents indicated their sex as female. For each visit, we identified whether the patient indicated their race(s) as (i) American Indian or Alaskan Native, (ii) Asian, (iii) Black or African American, (iv) Native Hawaiian or Other Pacific Islander or (v) White, and whether the patient indicated their ethnicity as of Hispanic or Latino or Spanish Origin. Additionally, we used the start date of the visit to categorize the visit by month between February 2020 and March 2022. We identified 16,220 distinct ICD10 clinical diagnosis codes listed for patients in the 26 month period not including U07.1, which was indicated for COVID-19 in October 2020 and generalized the diagnoses codes to 1,600 distinct category levels codes. All variables were treated as a binary categorical variables with 1 indicating that the patient was a part of the age group, or self-identified as female or self-identified as the specific race or ethnicity or or had a diagnosis code listed during that visit and 0 indicating the inverse.

### Training and evaluating the random forest classifier

We used Python 3.9.10 with sklearn 0.24.2 and pickle) libraries to fit, evaluate and apply a random forest model. The random forest classifier was refined using maximum depth and the number of estimators to maximize AUROC in the independent evaluation set.

### Identifying phenotypes associated with COVID-19

Clinical diagnosis data from each visit between February 2020 and March 2022 were mapped from the ICD10 vocabulary to PheCodes. Additionally, historical condition data from our clinical data warehouse was mapped from SNOMED vocabulary to PheCodes. We used Python 3.9.10 with numpy 1.19.5, pandas 1.2.3, and scipy 1.6.2 libraries to statistically evaluate the distributions. For visits with a follow up within each time interval (e.g. within 1 week), we discerned the visits where the PheCode was observed in the followup and the visits where the PheCode was not observed and compared between the distributions using a Mann-Whitney U test. *p*-values of 0 are presented as *p* < 2.225E-308 (the minimum value for a float object in Python) in the manuscript and tables, while *p*-values of 0 are recast as half the minimum non-zero *p*-value per test for stylistic purposes in figures. In evaluating instances where the patient was not previously diagnosed with the condition, we eliminated all patients who had a previous history of the condition (i.e. had the diagnosis prior to the start of the visit).

### Cox Proportional Hazards modeling and Kaplan-Meier curve fitting

From our cases visits (those visits where the patient returned with the condition within one year), we identified the time to event as the time from the end of the preceding visit to the the first instance of the condition within one year of the visit. In our non-case visits, we censored the data at the final interaction with NYP/CUIMC within the time period. We used Python 3.9.10 with numpy 1.19.5, pandas 1.2.3, and lifelines 0.25.10 libraries to determine and statistically evaluate the hazards ratios associated with COVID-19 probability. In order to build Kaplan-Meier curves, we stratified our data by the COVID-19 probability of the preceding visit (≤ 0.2. > 0.2 and ≤ 0.4, > 0.4 and ≤ 0.6, > 0.6 and ≤ 0.8, and > 0.8) and fit individual curves to each stratified dataset.

### Data availability

All supplementary tables are available from GitHub as .csv files (https://github.com/tatonetti-lab/predict-covid-effects).

### Code availability

All scripts used for data preparation and analysis are available from GitHub as Jupyter Notebooks (https://github.com/tatonetti-lab/predict-covid-effects).

## Data Availability

Data availability
All supplementary tables are available from GitHub as .csv files (https://github.com/tatonetti-lab/predict-covid-effects).
Code availability
All scripts used for data preparation and analysis are available from GitHub as Jupyter Notebooks (https://github.com/tatonetti-lab/predict-covid-effects).

